# Effect of anakinra on mortality in COVID-19: a patient level meta-analysis

**DOI:** 10.1101/2021.04.13.21255411

**Authors:** Evdoxia Kyriazopoulou, Thomas Huet, Giulio Cavalli, Andrea Gori, Miltiades Kyprianou, Peter Pickkers, Jesper Eugen-Olsen, Mario Clerici, Francisco Veas, Gilles Chatellier, Gilles Kaplanski, Mihai G. Netea, Emanuele Pontali, Marco Gattorno, Raphael Cauchois, Emma Kooistra, Matthijs Kox, Alessandra Bandera, Hélène Beaussier, Davide Mangioni, Lorenzo Dagna, Jos W. M. van der Meer, Evangelos J Giamarellos-Bourboulis, Gilles Hayem, International collaborative group for Anakinra in COVID-19

## Abstract

**Background:** Anakinra may represent an important therapy to improve the prognosis of COVID-19 patients. This meta-analysis using individual patient data was designed to assess the efficacy and safety of anakinra treatment in patients with COVID-19.

**Methods:** Based on a pre-specified protocol (PROSPERO: CRD42020221491), a systematic literature search was performed in MEDLINE (PubMed), Cochrane, medRxiv.org, bioRxiv.org and clinicaltrials.gov databases for trials in COVID-19 comparing administration of anakinra with standard-of-care and/or placebo. Individual patient data from eligible trials were requested. The primary endpoint was the mortality rate and the secondary endpoint was safety.

**Findings:** Literature search yielded 209 articles, of which 178 articles fulfilled screening criteria and were full-text assessed. Aggregate data on 1185 patients from 9 studies were analyzed and individual patient data on 895 patients from 6 studies were collected. Most studies used historical controls. Mortality was significantly lower in anakinra-treated patients (38/342 [11·1%]) as compared with 137/553 (24·8%) observed in patients receiving standard-of-care and/or placebo on top of standard-of-care (137/553 [24·8%]); adjusted odds ratio (OR), 0·32; 95% CI, 0·20 to 0·51; p <0·001. The mortality benefit was similar across subgroups regardless of diabetes mellitus, ferritin concentrations, or baseline P/F ratio. The effect was more profound in patients exhibiting CRP levels >100 mg/L (OR 0·28,95%CI 0·27-1·47). Safety issues, such as increase of secondary infections, did not emerge.

**Interpretation:** Anakinra may be a safe anti-inflammatory treatment option in patients hospitalized with moderate-to-severe COVID-19 pneumonia to reduce mortality, especially in the presence of hyperinflammation signs such as CRP>100mg /L.

**Funding:** Sobi.

**Research in context:** *Evidence before this study:* Since the emergence of the COVID-19 pandemic, numerous drugs have been tried in an effort to prevent major detrimental consequences, such as respiratory and multiorgan failure and death. Early during the pandemic, it was realized that drugs aiming to regulate the immune host reaction may play an important role in the treatment of COVID-19. Evidence from a small number of patients with moderate or severe COVID-19 treated with anakinra, and interleukin-1 receptor antagonist, has suggested therapeutic efficacy. We systematically searched all available literature and aimed to present cumulative evidence of anakinra treatment in COVID-19 and the related effect on mortality.

*Added value of this study:* This is the first patient-level analysis on the effect of anakinra treatment in COVID-19 patients, which, on the one hand, suggests a significant benefit in the reduction of mortality and on the other hand, reassures safety of the treatment. Most importantly, the current study identifies a subgroup of patients with CRP>100mg/L, that may benefit most from treatment with anakinra. Confirmation of these effects in larger randomized clinical trials (RCTs) is urgently needed.

*Implications of all the available evidence:* Anakinra may be an effective and safe immunomodulatory treatment in moderate-to-severe cases of pneumonia due to COVID-19 to prevent unfavorable outcomes. Anakinra may be helpful to avoid adverse events, such as breakthrough infections observed often with dexamethasone use, and may be considered an alternative in specific subgroups of patients e.g. diabetics. Larger trials, summarized in the Table, are ongoing and their results are urgently needed to investigate anakinra’s best place in the treatment of COVID-19. 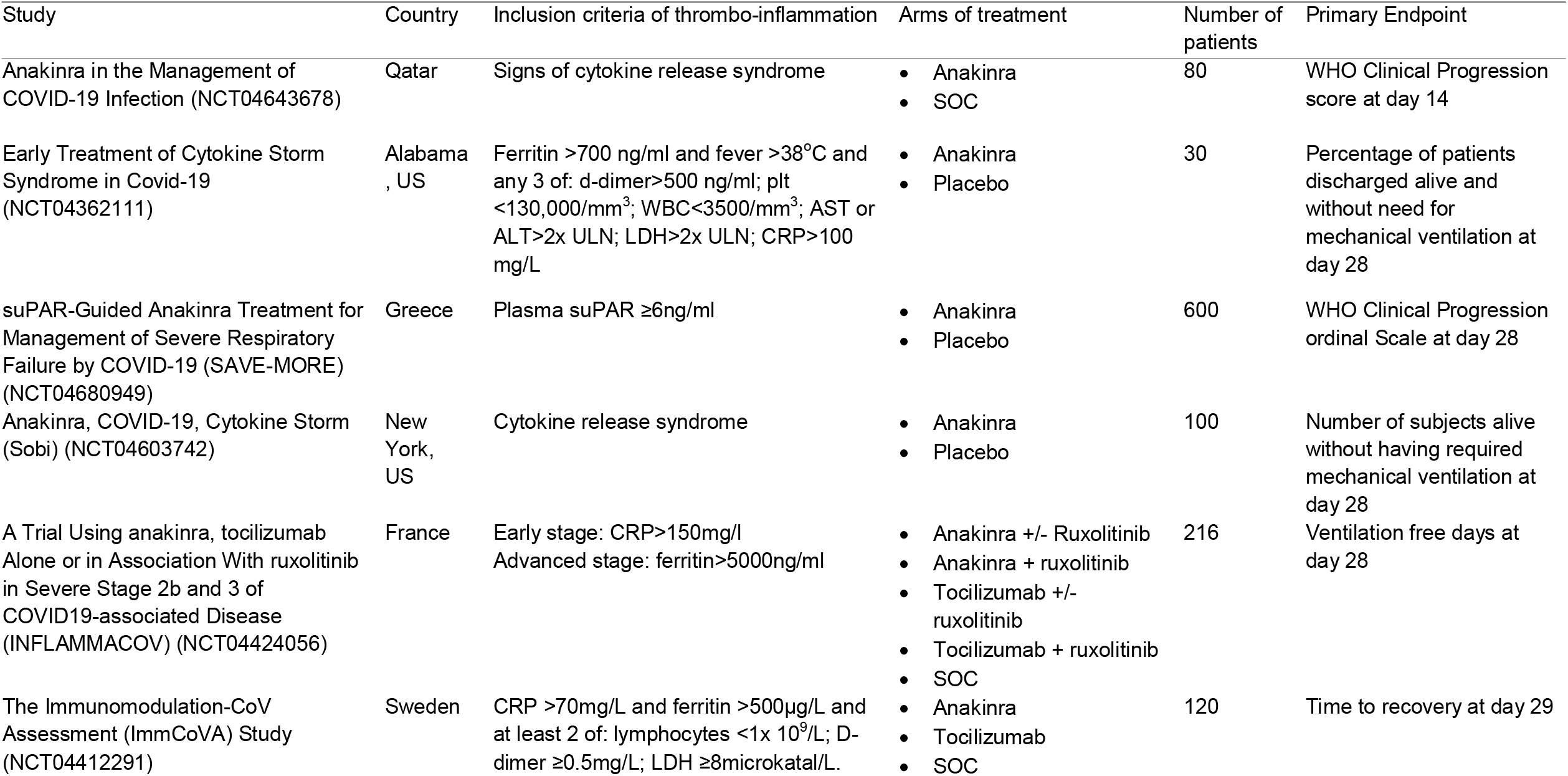

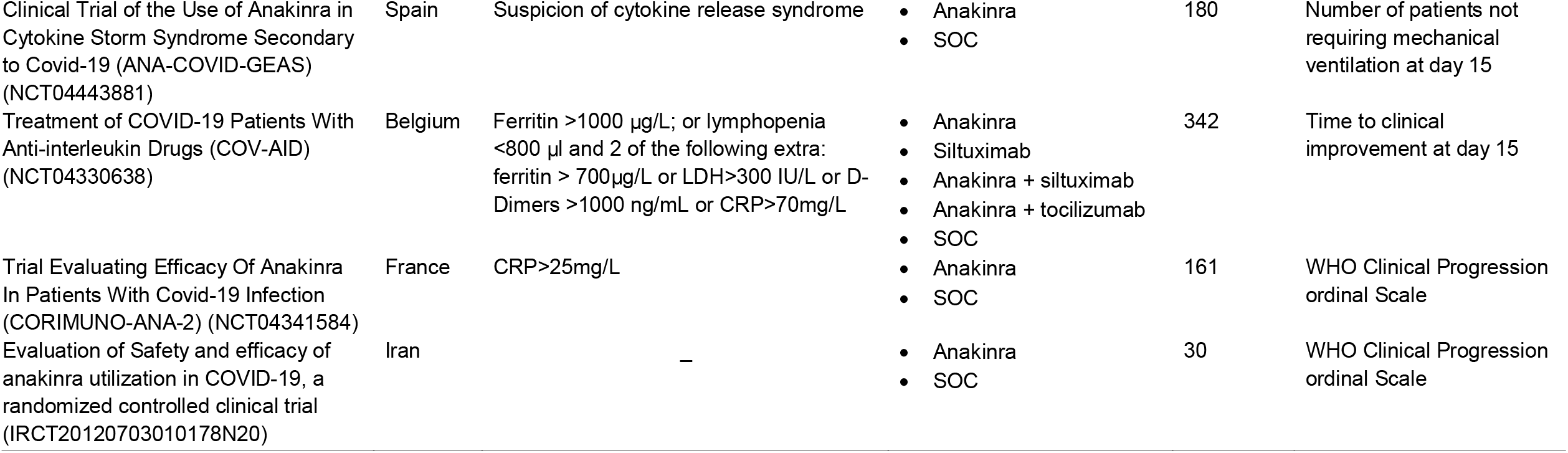

## Introduction

COVID-19 is caused by the severe acute respiratory syndrome coronavirus (SARS-CoV)-2 and is recognized as a pandemic, affecting more than 200 countries with millions of confirmed cases and deaths worldwide (1). The most severe and lethal forms of COVID-19 have been linked to different and possibly combined physiopathological processes, involving severe acute diffuse alveolar damage and hyaline deposits in the lungs induced by SARS-CoV-2, multiple arterial and venous thromboses resulting from both endothelitis and hypercoagulability, and a hyperinflammatory response with overproduction of pro-inflammatory cytokines. Although systemically, there may not be a “cytokine storm” (2-4), there are clear indications of pulmonary compartmentalization of hyperinflammation (5-7).

In order to control the SARS-CoV-2-induced hyperinflammatory syndrome, during which the IL-1/IL-6 pathway is mainly involved, various targeted therapies have been tested, predominantly in combination with antibiotics and anticoagulants. Anakinra, a recombinant interleukin (IL)-1 receptor antagonist, early on appeared as a logical candidate, since it had previously been tested with encouraging results in other hyperinflammatory situations, including the macrophage activation syndrome complicating severe bacterial sepsis and the cytokine release syndrome observed during antitumoral chimeric antigen receptor (CAR)-T cell therapy (8-10).

Case series of a small number of patients with moderate or severe COVID-19 treated with anakinra have been published early during the pandemic suggesting promising results and paving the way for additional larger studies (11-14). In most studies, anakinra was administered as an off-label use in patients with clinical and/or laboratory signs of hyperinflammation or in a context of clinical trials, in an attempt to dampen the IL-1 driven inflammation which is believed to play a significant role in COVID-19 pathophysiology. In this systematic review and meta-analysis, we aimed to present all available cumulative evidence on anakinra treatment in COVID-19, to investigate the effect of anakinra on mortality rate, and to better characterize the candidate population most likely to benefit from such a tailored immunotherapy.

## Methods

### Search strategy and selection criteria

This meta-analysis was conducted according to the Preferred Reporting Items for Systematic reviews and Meta-Analyses (PRISMA) statement (15), based on a pre-specified protocol (PROSPERO: CRD42020221491). Eligibility criteria were defined using the PICO statement as follows; P: hospitalized patients with confirmed COVID-19. I: treatment with anakinra. C: patients receiving standard-of-care and/or placebo comparator on top of standard-of-care, whereas O was the rate of mortality, and the rate of common adverse events such as elevated liver function tests, leukopenia and secondary infections. All randomized trials, comparative studies and observational studies, published as full text in English were included, while editorials, conference abstracts, animal studies, case reports, articles not written in English language or not providing full text were excluded. Systematic reviews were consulted for additional information, but were excluded to avoid duplication.

Search was conducted on December 28, 2020 and repeated on January 22, 2021 by two independent authors (E.K. and E.J.G.B) across MEDLINE (PubMed), Cochrane, medRxiv.org, biorxiv.org and clinicaltrials.gov databases using the following terms: “COVID-19” or “SARS-CoV-2” and “anakinra”, “interleukin-1”, “interleukin blockade”. Detailed strategy is provided in Supplement. Both reviewers assessed all articles first on the title, then on the abstract and finally the full text to identify those eligible. Any between-reviewer controversies were resolved by discussion. Individual patient data (IPD) were requested from investigators/corresponding authors of all eligible articles. Data from each article was first checked against reported results and queries were resolved with the investigators. Data has been assessed in a consistent manner across all trials with standard definitions and parameters. The following variables were collected: first author name, country of origin, publication time, study design, total number of patients, criteria for enrollment, number of patients treated with anakinra, number of controls, age and comorbidities of participants, serum ferritin and C-reactive protein levels at baseline, respiratory ratio at baseline, route of anakinra administration, intake of steroids, mortality, onset of respiratory failure necessitating invasive mechanical ventilation, onset of adverse events (elevated liver function tests, leukopenia, breakthrough infections). If IPD were not available, all above available data were extracted independently by the two authors from the original publication.

The quality of each study was evaluated by the 2 reviewers with the Newcastle-Ottawa scale (16). Along the entire process, between-reviewer controversies were resolved by discussion.

### Statistical methods

The primary endpoint was the mortality rate. Secondary endpoint was safety, assessed by the rate of adverse events. Odds ratios and their respective 95% confidence intervals (CI) were the effect measure used for comparing anakinra-treated patients to control groups for each end-point. Pre-defined subgroup analyses were performed for the primary endpoint according to the following variables: baseline respiratory ratio, baseline CRP and serum ferritin levels and Charlson’s comorbidity index. First, a logistic regression analysis was used to detect any interaction effect of anakinra treatment with the different studies and second, to detect if anakinra effect on primary outcome remains independent of factors interfering with mortality (age, Charlson’s comorbidity index, CRP and P/F ratio calculated as the ratio of the arterial partial oxygen pressure divided by the fraction of inspired oxygen). Already described cutoffs were used for the characterization of above the subgroups (17-20). Sensitivity analyses were planned on i) time period (before and after implementation of dexamethasone) and ii) RCTs. Aggregate data were meta-analyzed by the Mantel-Haenszel method. The Sidik-Jonkman estimator was used for tau^2^ and the Q-profile method for confidence interval of tau^2^ and tau. Continuity correction of 0.1 was applied in studies with zero cell frequencies. Finally, heterogeneity and inconsistency were assessed in a meta-analysis of aggregate data from all eligible trials using the I^2^ criterion. For I^2^ <50% the fixed effects model was used while for I^2^ ≥ 50% the random effects model. The corresponding forest plot was produced, while publication bias was assessed with the funnel plot. Trials without available IPD participated only in the aggregate data meta-analysis. No imputation for missing data within the IPD was made. All statistical analyses were done using SPSS (version 23) and Review Manager (version 5.3).

## Results

Initial search yielded 209 articles; after removal of duplicates and articles with irrelevant titles, 178 were screened full-text by the reviewers. After applying exclusion criteria, 9 studies, for which aggregate data were available, including a total of 1185 patients were finally analyzed, and IPD were available for 6 of them including a total of 895 patients (Figure 1). A total of 103/895 patients were under mechanical ventilation at baseline. Among the three studies for which IPD were not obtained, two favored and one did not favor anakinra treatment. Characteristics of the included trials are presented in Table 1. Three studies were conducted in France (21-23), three in Italy (24-26), one in the Netherlands (27), one in Greece (28), and one in Oman (29). Data published early during the pandemic in an Italian study (30) were integrally included in a larger study of the same group (25): thus, to avoid duplication, the first cohort was not included. Seven studies were conducted before dexamethasone implementation in the standard-of-care treatment of COVID-19 patients: one study recruited anakinra-treated patients after dexamethasone implementation but controls were recruited prior to its implementation, and only one study prospectively recruited both anakinra-treated and control subjects after implementation of dexamethasone. Assessment of individual risk of bias according to Newcastle-Ottawa scale is provided in eTable 1. The overall quality of included studies was high.

**Figure 1.**
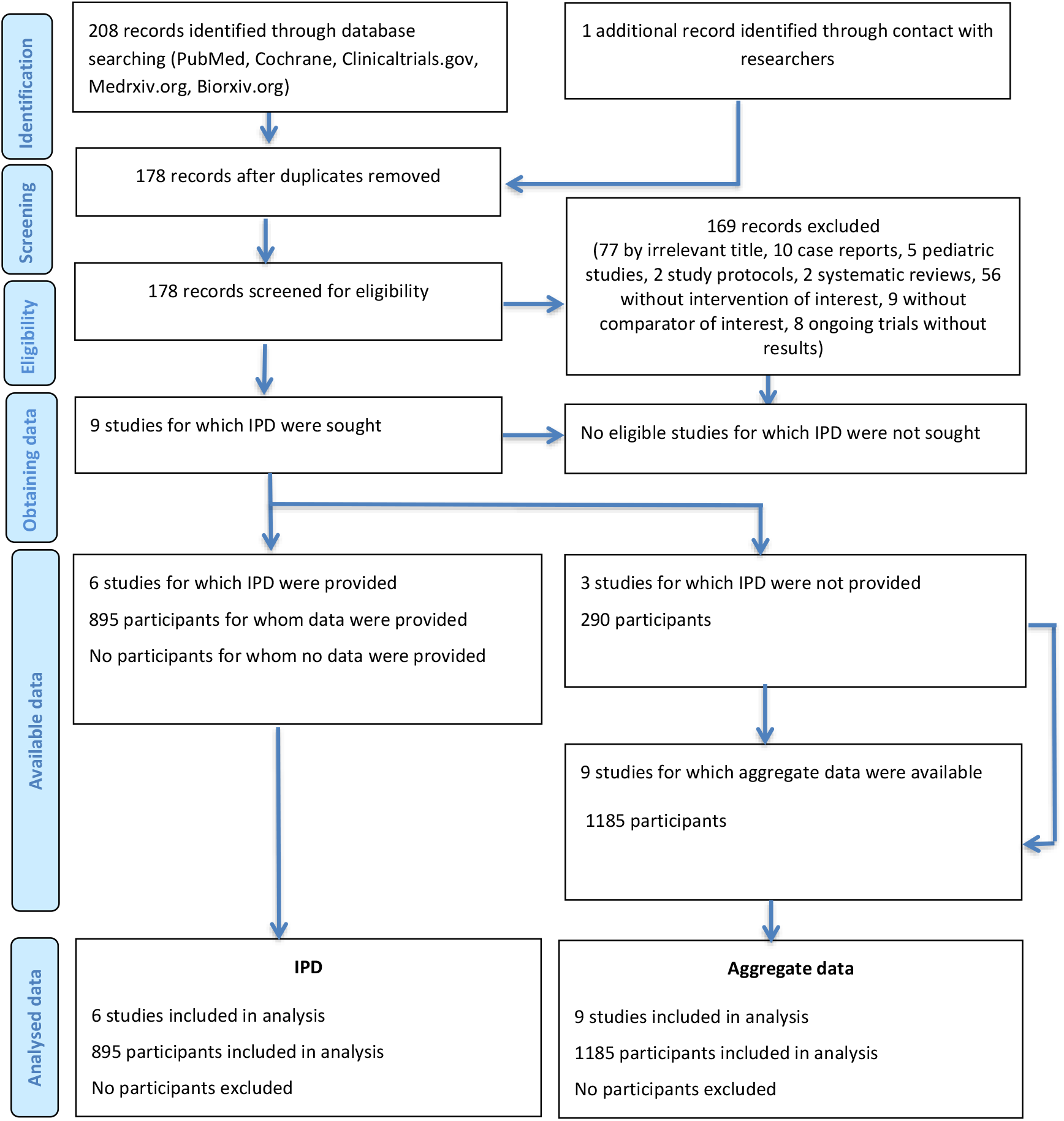
Study selection. Abbreviation: IPD, individual patient data.

**Table 1.**
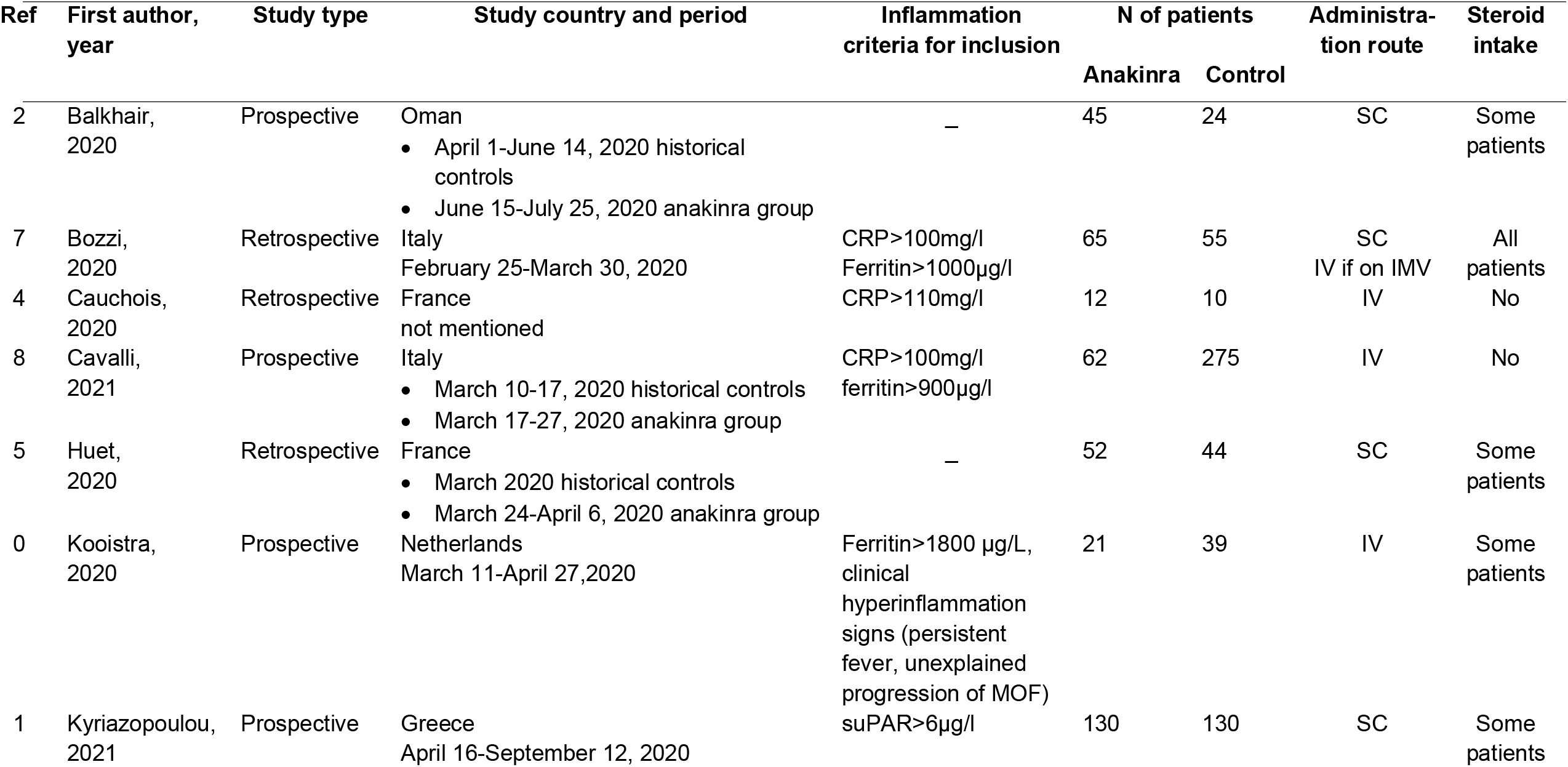
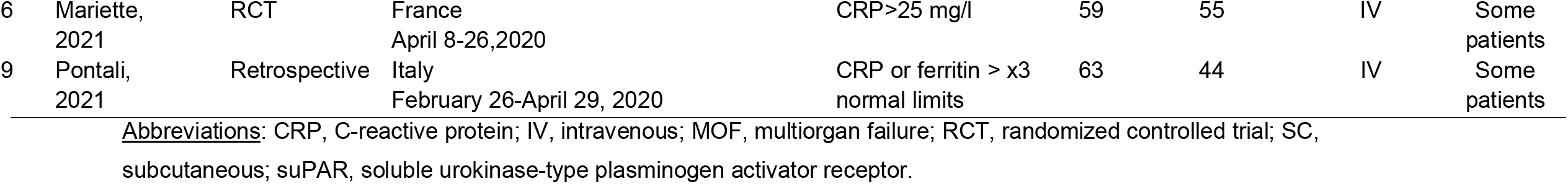
Characteristics of included studies.

Aggregate data from nine studies (1185 patients) support a significant reduction in mortality among anakinra-treated patients compared to controls receiving standard-of-care and/or placebo on top of standard-of-care; the pooled OR for mortality was estimated 0·37 (95% CI, 0·27-0·51; I^2^, 31%), as shown in Figure 2. There was no evidence of publication bias (test for funnel plot asymmetry: t = - 0·9137, p: 0·391; eFigure 1). In IPD analysis (895 patients), a total of 38 (11·1%) anakinra-tobreated patients died compared to 137 (24·8%) of comparators receiving standard-of-care and/or placebo on top of standard-of-care (OR, 0·38; 95% CI, 0·26 to 0·56; p: <0·001) (Table 2). No interaction effect was observed among the six different studies included and anakinra-treatment regarding the primary outcome (p: 0·147) (Table 2). After adjustment of age, comorbidities, baseline respiratory ratio, lymphopenia and elevated CRP levels, anakinra was proven an independent protective factor from mortality (adjusted OR, 0·32; 95% CI, 0·20 to 0·51; p <0·001) (Table 2). Similar OR were provided after adjustment for ferritin (available for 486 patients) and IL-6 levels (available for 530 patients). The predefined sensitivity analyses on RCTs were not possible to conduct, since only 1 RCT was available.

**Figure 2.**
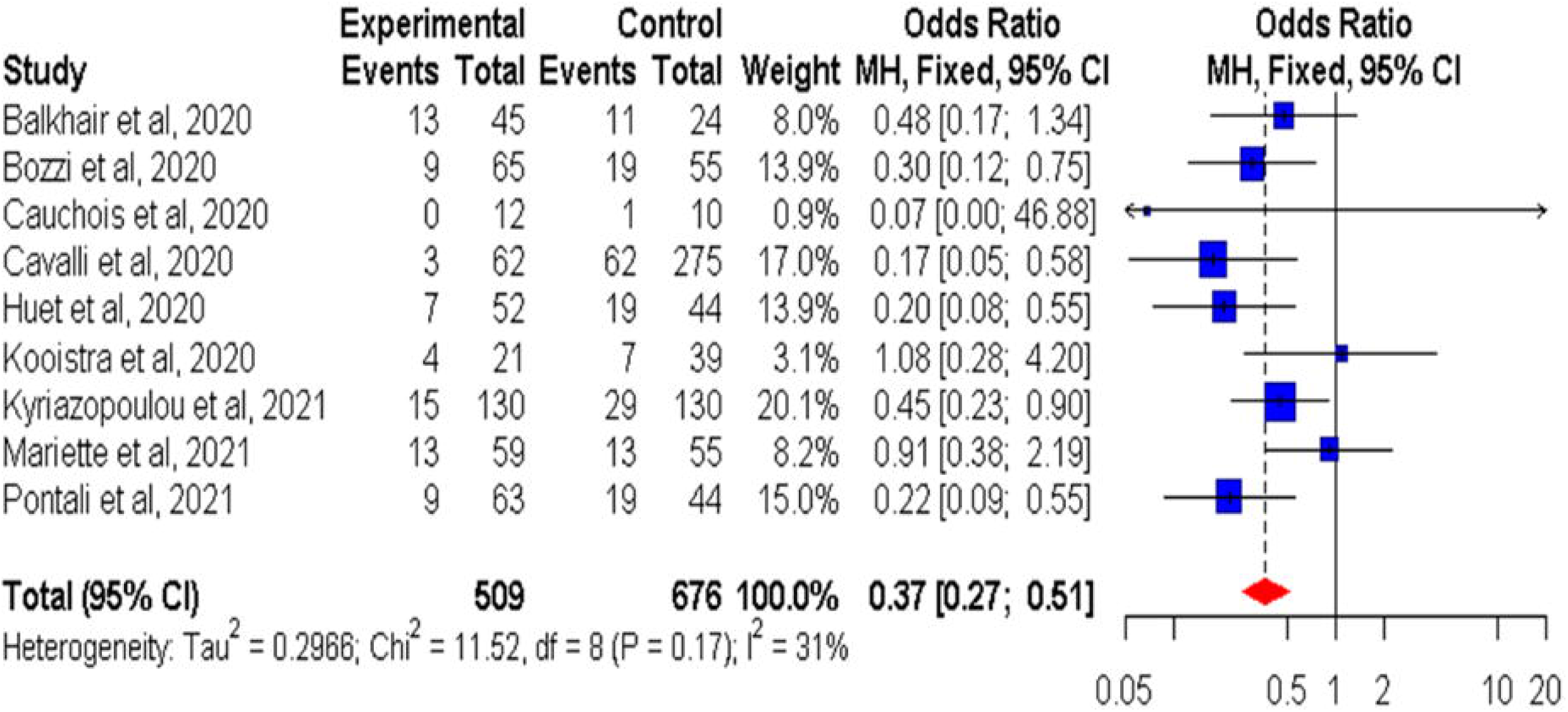
Forest plot showing mortality from aggregate data meta-analysis Odds ratios calculated with a fixed-effects Mantel-Haenszel test. Abbreviation: CI, confidence interval.

**Table 2.**
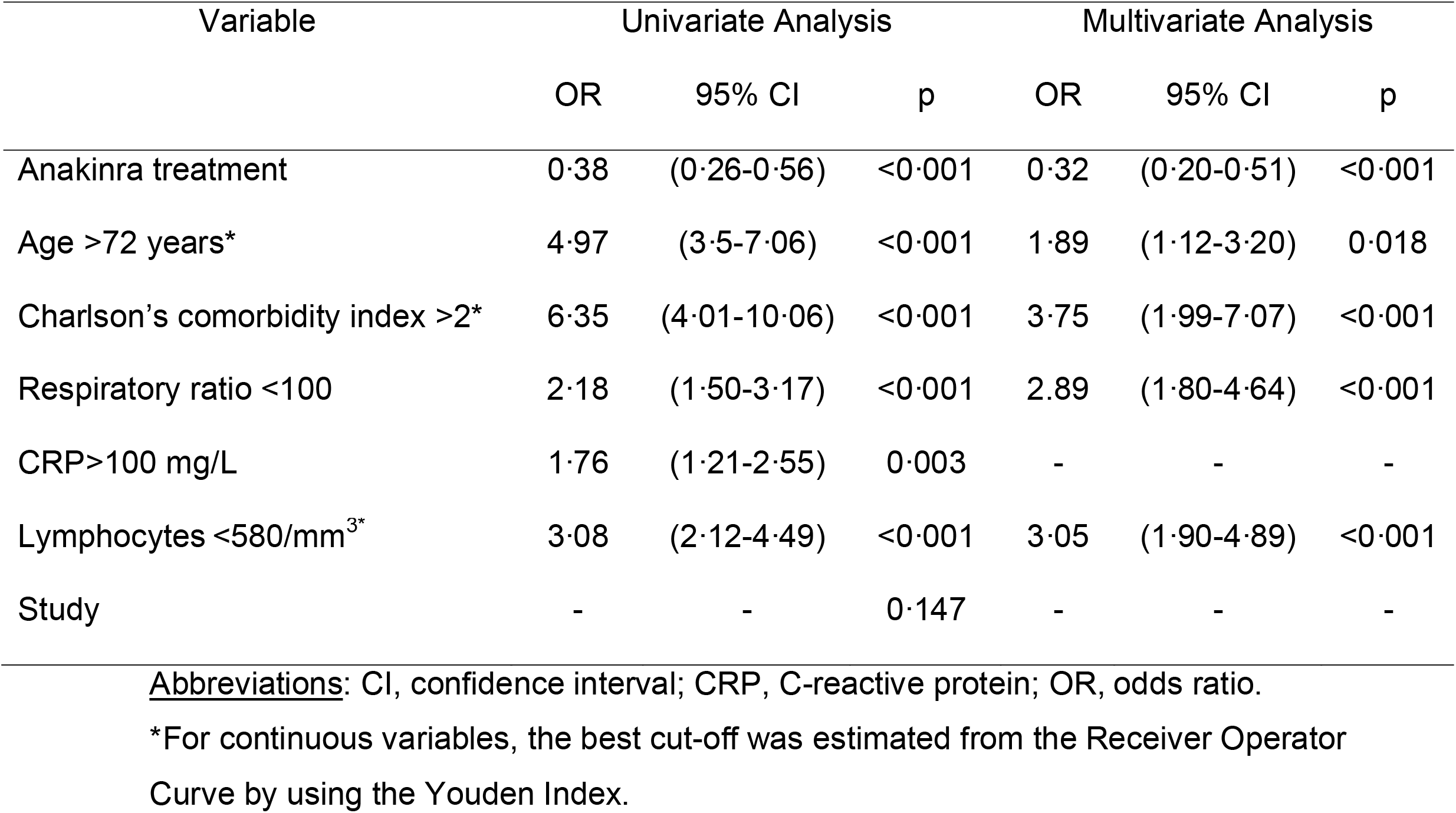
Univariate and multivariate logistic regression analysis of variables associated with mortality in the individual patient data analysis of 895 patients.

Anakinra treatment was significantly beneficial in absence of dexamethasone co-administration (n=559) (OR 0·23 95% CI 0·12-0·43); this was no longer the case by co-administration of dexamethasone (n=239) (OR 0·72, 95%CI 0·37-1·41; p_Breslow_ 0·012). Subgroup analysis per CRP and ferritin levels as well as baseline P/F ratio, showed that anakinra was more effective in lowering mortality in those patients presenting with CRP levels above 100 mg/L (OR 0·28,95%CI 0·27-1·47), but therapeutic efficacy appeared not related to ferritin level, or P/F ratio (Figure 3). Anakinra effect on mortality was similar in patients (n=116) with (OR 0·40; 95% CI 0·17-0·91) and without (n=299) diabetes mellitus (OR 0·37, 95% CI 0·19-0·74; p_Breslow_ 0·902).

**Figure 3.**
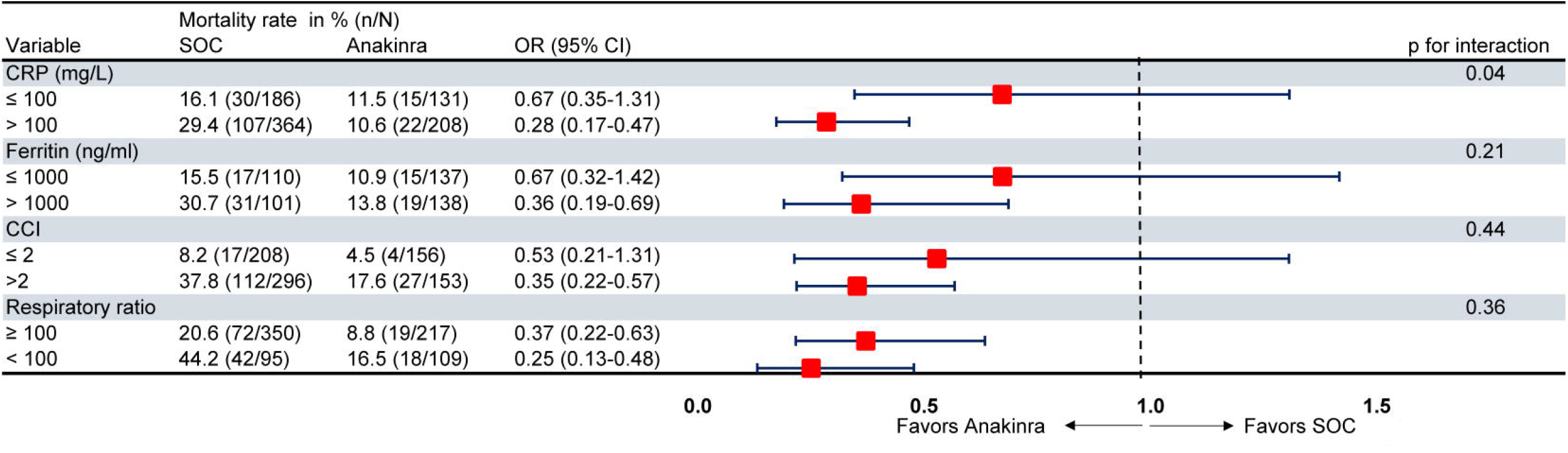
Subgroup analysis of mortality between anakinra-treated patients versus comparators. P values of the interaction effect of the treatment on mortality, in each subgroup and among the studies are provided. Abbreviations: CCI Charlson’s comorbidity index; CI, confidence interval; CRP, C-reactive protein; OR, odds ratio; SOC, standard of care

Safety of anakinra was investigated as a secondary endpoint. The pooled OR of elevation of liver function tests with anakinra treatment was 3·00 (95% CI, 0·26 to 34·66, I^2^, 85%) (eFigure 2A) and the respective OR for onset of leukopenia and secondary infection was 3·71 (95% CI, 0·49 to 27·84, I^2^, 51%) (eFigure 2B) and 1·35 (95% CI, 0·59 to 3·10, I^2^, 79%) (eFigure 2C), respectively. Thromboembolic events were reported in only two studies (23,28); thus, no meta-analysis on this endpoint was performed. Nevertheless, in both studies, anakinra did not increase the thromboembolic risk compared to control.

## Discussion

This systematic review and meta-analysis of the available published studies argues that treatment with anakinra reduces mortality in hospitalized patients with pneumonia due to COVID-19 when compared with patients receiving standard-of-care and/or placebo on top of standard-of-care. This benefit on survival was most profound in hyper-inflamed patients with CRP levels >100mg/L. Anakinra treatment did not raise safety concerns in the study population.

The first two cohort studies dealing with anakinra in severe forms of COVID-19 were not randomized and used patients receiving usual care in the same institution as historical controls (22,30). Although the design clearly has its limitations, as general care for COVID-19 patients improved over time, possibly regardless of the anakinra treatment, the observed magnitude of protective effect of anakinra was convincing enough to continue with additional studies that globally supported a positive effect of anakinra in severe forms of COVID-19. The open-label single-arm phase II prospective SAVE trial concluded not only that suPAR-guided anakinra treatment prevented onset of respiratory failure necessitating mechanical ventilation but also reduced both short (30-day)- and long (90-day)-mortality compared with standard-of-care alone (28). suPAR is an early predictor of some of the most critical COVID-19 outcomes, including respiratory failure, kidney failure and mortality (31,32). The only randomized controlled trial published so far was initiated by the French consortium CORIMUNO, but was prematurely interrupted following enrolment of 59 anakinra and 55 usual care patients because of assumed futility (23). The results from the CORIMUNO-ANA-1 study suggest that anakinra was not effective in reducing the need for non-invasive or mechanical ventilation or death in patients with COVID-19 and mild-to-moderate pneumonia. However, in the CORIMUNO-ANA-1 study, the day-14 mortality was 15% in the anakinra group compared to 24% in the usual care group and the WHO Clinical Progression Scale (CPS) was also suggestive for a beneficial effect of anakinra at day 14. Unfortunately, stopping for futility increases the risk of imbalance in prognostic factors and leaves the primary research question unanswered. This is why stopping any trial early is controversial, especially when the decision for early stopping does not follow a pre-specified stopping boundary. In addition, the cut-off criterion chosen for CRP was relatively low (>25 mg/L), which suggests that a proportion of patients had not yet entered the hyperinflammatory phase of COVID-19, during which anakinra is probably the most effective (33). Results from the CORIMUNO-ANA-2 study in patients with more severe COVID-19, hospitalized in the ICU, are still awaited.

Predicting unfavorable outcome during COVID-19 is a real challenge. Algorithms have been constructed and validated, taking into account a number of clinical variables, including older age, male sex, high body mass index and comorbidities (34-36). Marked lymphopenia, decreased CD3, CD4, or CD8 T-lymphocyte counts, thrombocytopenia, and markedly elevated inflammatory biomarkers were associated with severe disease and the risk of developing sepsis and rapid unfavorable disease progression. Accumulating evidence suggests that some patients with severe COVID-19 suffer from a hyperinflammatory phenotype described as “cytokine storm”. Cytokine Storm Syndrome (CSS) is a life-threating condition requiring intensive care and resulting in a quite high mortality. It is characterized by an overwhelming systemic inflammation, with hyperferritinemia, hemodynamic instability, and multi-organ failure. The trigger for CSS is suggested to be extensive cell death (i.e. induced by viral replication), which prompts massive release of pro-inflammatory intracellular mediators and danger signals, notably including IL-1α (37). These in turn induce an uncontrolled immune response resulting in continuous activation and expansion of immune cells, lymphocytes, and macrophages, which produce large amounts of proinflammatory cytokines like IL-1β, IL-6, IL-18, as well as IFN-γ and TNF-α (38). Clearly not all COVID-19 patients experience a cytokine-storm (7), but even in these patients the pulmonary inflammatory response may be exacerbated and they may benefit from immunomodulating therapies (5,6).

While the molecular mechanisms driving disease severity remain unclear, the clinical association of inflammatory mediators such as IL-6 with severe cases suggests that excessive inflammation plays a fundamental role in determining a bad clinical outcome (39-40). The induction of IL-1-dependent inflammatory processes in host cells mainly requires the engagement of inflammasomes, which are protein platforms that aggregate in the cytosol in response to various stimuli (41-42). The presence of cell death and inflammasome-derived products such as IL-1β, IL-18, and LDH in patients’ sera suggests the implication of inflammasome (43-45). However, these compounds can also be produced *via* inflammasome-independent pathways.

SARS-CoV-2 was shown to stimulate the NLRP3 pathway, inducing an inflammasome-dependent overproduction of IL-1, a pivotal phenomenon resulting in the already mentioned “cytokine storm”, involving various other pro-inflammatory mediators, including IL-6. In the inflammasome-dependent cascade, IL-1 is located upstream of IL-6, as well other inflammatory mediators (43-48). As a proof, decrease in serum IL-6 concentration classically follows the effective inhibition of IL-1. Such a hierarchical process could explain why targeting IL-1 appears more efficient in hyperinflammatory forms of COVID-19, than IL-6 inhibition.

IL-6 is a potent stimulus of production of CRP by the liver (49). Therefore, CRP levels probably represents the simplest biological test to indirectly evaluate the intensity of the ongoing cytokine storm. Indeed, CRP titers above 100 mg/L have been associated with a worse prognosis in severe forms of COVID-19. It could also represent a signal indicating a window of opportunity for using targeted immune-active drugs, aiming to inhibit the inflammasome pathway and the resulting overproduction of pro-inflammatory cytokines. Our results underline a profound benefit of anakinra in the subgroup of patients with CRP levels >100mg/L.

Anakinra did not appear to raise the risk for secondary infection in our study and may therefore represent a safer alternative to dexamethasone with similar, if not better, effect on final outcome. An additional benefit of anakinra may be seen in special populations, such as diabetics, that may be more susceptible for secondary infections (50). Indeed, our study shows similar benefit of anakinra in the subgroup of patients with diabetes mellitus.

To the best of our knowledge, this is the first patient-level meta-analysis on the effect of anakinra treatment in COVID-19, which first, outscores a significant benefit in the reduction of mortality and second, characterizes a subgroup of patients with CRP>100mg/L as the best candidate for this treatment. The retrospective observational character of the majority of the studies included, as well as the fact that most of them were performed before the implementation of dexamethasone as standard-of-care are clearly limitations of the current analysis. A randomized placebo-control trial with the acronym SAVE-MORE (NCT04680949) is ongoing in Greece and Italy comparing anakinra to placebo in patients receiving already standard-of-care in relation to 28-day outcome, as expressed in the 11-point WHO CPS scale and anakinra is also one of the interventions tested in REMAP-CAP. To conclude, anakinra treatment in patients hospitalized with pneumonia due to COVID-19 appears to be an efficient option with favorable safety profile and may reduce mortality in moderate-to-severe cases, especially if patients present with hyperinflammation signs such as CRP>100mg/L. Larger randomized trials are urgently needed to provide definitive conclusions regarding the place of anakinra in the anti-COVID19 armamentarium.

## Data Availability

All available data are included in manuscript and supplement.

## Contribution

E.Ky. performed literature search and study selection, participated in data analysis and drafted the manuscript. E.J.G.B. conceptualized the study, participated in literature search and study selection. M.Ky. performed data analysis. G.H drafted the manuscript. T.H., G.Ca., A. G., P.P., G.Ch., G.K., E.P., M.G., R.C., E.Ko, M.K., H.B., D.M., A.B. and L.D. provided clinical data and revised the manuscript for important intellectual content. J.E.O, M.C., F.V., M.G.N and J.W.M. revised the manuscript for important intellectual content. All authors gave approval for the version to be published.

