## Supplement for "Effect of anakinra on mortality in COVID-19: a patient level meta-analysis"

**“Online Supplement”**

**Search Strategy.**

| **PubMed (Date Run: 22/01/2021)** | | |
| --- | --- | --- |
| Step | **Search strategy** | **Found** |
| **#1** | (((COVID-19[MeSH Terms]) AND (anakinra[MeSH Terms])) OR ((COVID-19[MeSH Terms]) AND (interleukin blockade[MeSH Terms])) OR ((COVID-19[MeSH Terms]) AND (interleukin-1[MeSH Terms])) OR ((SARS-CoV-2[MeSH Terms]) AND (anakinra[MeSH Terms])) OR ((SARS-CoV-2[MeSH Terms]) AND (interleukin blockade[MeSH Terms])) OR ((SARS-CoV-2[MeSH Terms]) AND (interleukin-1[MeSH Terms]))) | 95 |
| **#2** | (((COVID-19[MeSH Terms]) AND (anakinra[MeSH Terms])) OR ((COVID-19[MeSH Terms]) AND (interleukin blockade[MeSH Terms])) OR ((COVID-19[MeSH Terms]) AND (interleukin-1[MeSH Terms])) OR ((SARS-CoV-2[MeSH Terms]) AND (anakinra[MeSH Terms])) OR ((SARS-CoV-2[MeSH Terms]) AND (interleukin blockade[MeSH Terms])) OR ((SARS-CoV-2[MeSH Terms]) AND (interleukin-1[MeSH Terms]))) Filters: Humans, English | 92 |

| **Cochrane Central Register of Clinical Trials (Date Run: 22/01/2021)** | | |
| --- | --- | --- |
| ID | **Search strategy** | **Found** |
| **#1** | (COVID-19):ti,ab,kw OR ("SARS Co-V"):ti,ab,kw | 3728 |
| **#2** | ("anakinra"):ti,ab,kw OR ("interleukin-1 blockade "):ti,ab,kw | 412 |
| **#3** | #1 AND #2 | 30 |
| **#4** | Removal of dublicates | 1 |

| **Medrxiv.org (Date Run: 22/01/2021)** | | |
| --- | --- | --- |
| ID | **Search strategy** | **Found** |
| **#1** | Anakinra AND COVID-19 | 10 |
| **#2** | Interleukin blockade AND COVID-19 | 6 |
| **#3** | Interleukin-1 AND COVID-19 | 7 |
| **#4** | (Anakinra OR interleukin blockade OR interleukin-1) AND COVID-19 | 23 |

| **Biorxiv.org (Date Run: 22/01/2021)** | | |
| --- | --- | --- |
| ID | **Search strategy** | **Found** |
| **#1** | anakinra AND COVID-19 | 4 |
| **#2** | Interleukin blockade AND COVID-19 | 0 |
| **#3** | Interleukin-1 AND COVID-19 | 22 |
| **#4** | (Anakinra OR interleukin blockade OR interleukin-1) AND COVID-19 | 26 |

| **Clinicaltrials.gov (Date Run: 22/01/2021)** | | |
| --- | --- | --- |
| ID | **Search strategy** | **Found** |
| **#1** | anakinra AND COVID-19 | 34 |
| **#2** | Removal of dublicates | 32 |

**eTable 1.** Risk assessment of included studies according to Newcastle-Ottawa Quality Assessment Scale.

|  | **Selection** | | | | **Comparability** | **Outcome** | | |  |
| --- | --- | --- | --- | --- | --- | --- | --- | --- | --- |
| **Ref.** | **Representativeness of the exposed cohort** | **Selection of the non-exposed cohort** | **Ascertainment of exposure** | **Demonstration that outcome of interest was not present at study start** | **Comparability of cohorts on the basis of the design or analysis** | **Assessment of outcome** | **Was follow-up long enough for outcomes to occur** | **Adequacy of follow up of cohorts** | **Total** |
| 25 | * | * | * | * | ** | * | * | * | 9 |
| 20 |  | * | * | * | ** | * | * | * | 8 |
| 17 |  | * | * | * | ** | * | * | * | 8 |
| 21 |  | * | * | * | ** | * | * | * | 8 |
| 18 | * | * | * | * | ** | * | * | * | 9 |
| 23 |  | * | * | * | ** | * | * | * | 8 |
| 24 |  | * | * | * | ** | * | * | * | 8 |
| 19 | * | * | * | * | ** | * | * | * | 9 |
| 22 | * | * | * | * | ** | * | * | * | 9 |

**Selection**

Representativeness of the exposed cohort (* for a and b) a) truly representative of the average in the community; b) somewhat representative of the average in the community; c) selected group of users eg nurses, volunteers; d) no description of the derivation of the cohort

Selection of the non-exposed cohort (* for a) a) drawn from the same community as the exposed cohort; b) drawn from a different source; c) no description of the derivation of the non-exposed cohort

Ascertainment of exposure (* for a and b) a) secure record (eg surgical records); b) structured interview; c) written self report; d) no description

Demonstration that outcome of interest was not present at start of study (* for a) a) yes; b) no

**Comparability**

Comparability of cohorts on the basis of the design or analysis a) study controls for the most important factor **; b) study controls for any additional factor*

**Outcome**

Assessment of outcome (* for a and b) a) independent blind assessment; b) record linkage; c) self report; d) no description

Was follow-up long enough for outcomes to occur (* for a) a) yes; b) no

Adequacy of follow up of cohorts (* for a and b) a) complete follow up - all subjects accounted for; b) subjects lost to follow up unlikely to introduce bias - small number lost; c) follow up rate large and no description of those lost; d) no statement

**eFigure 1.** Funnel plot of publication bias for trials included in the analysis of the primary endpoint. Test for funnel plot asymmetry: t = -0.9137, df = 7, p = 0.391

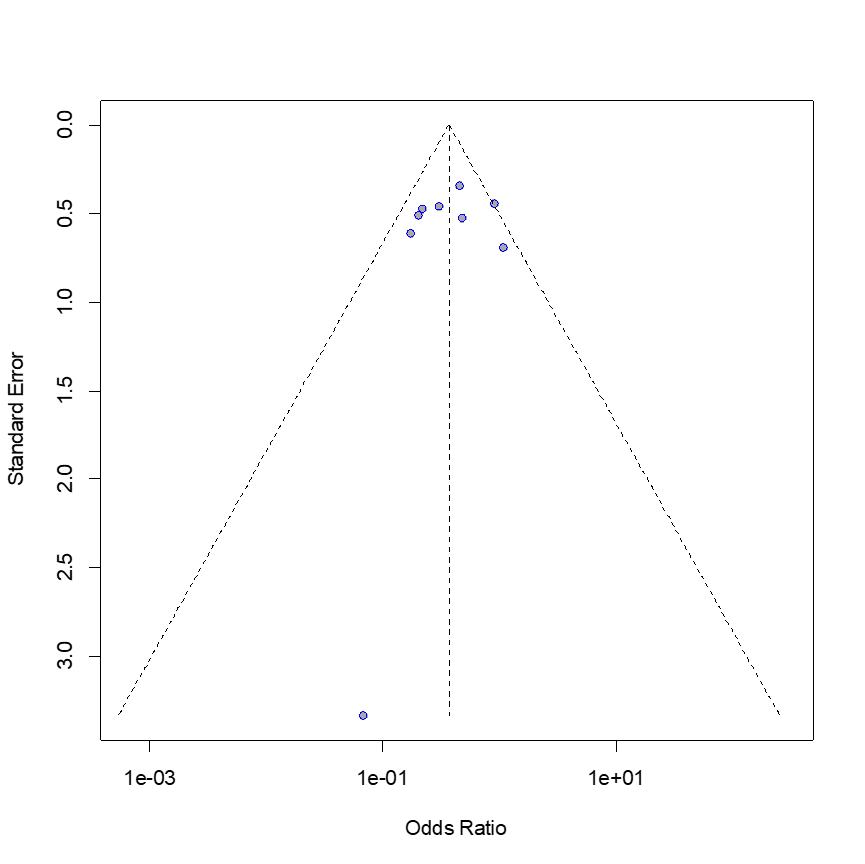

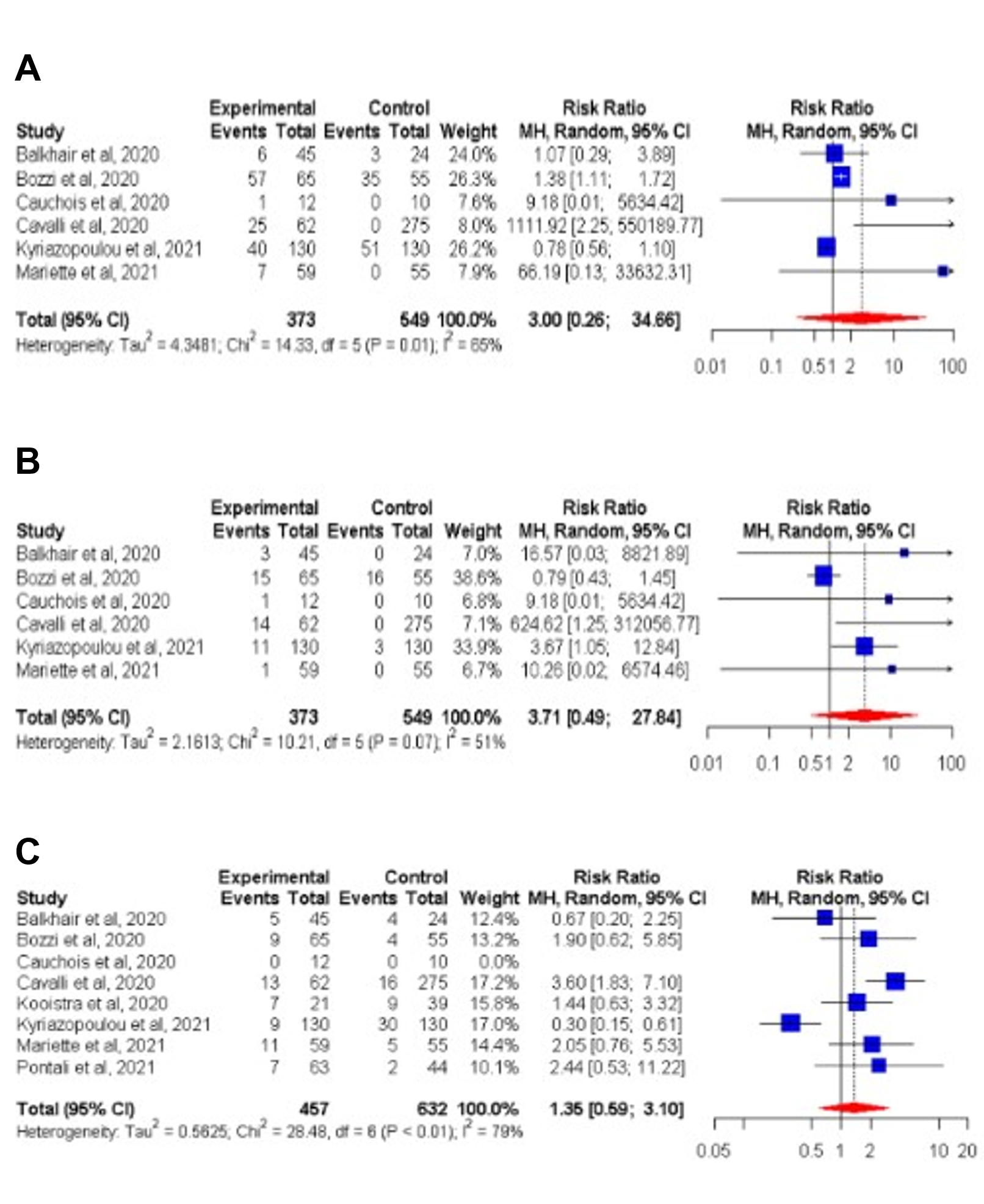
**eFigure 2.** Forest plot of pooled odds ratio ad respective confidence interval (CI) of onset of adverse events with anakinra treatment versus comparators. A) Elevated liver function tests; B) Leukopenia; and C) Onset of breakthrough infection.
